# IgG antibody seroconversion and the clinical progression of COVID-19 pneumonia: A retrospective, cohort study

**DOI:** 10.1101/2020.07.16.20154088

**Authors:** Kazuyoshi Kurashima, Naho Kagiyama, Takashi Ishiguro, Yotaro Takaku, Hiromi Nakajima, Shun Shibata, Yuma Matsui, Kenji Takano, Taisuke Isono, Takashi Nishida, Eriko Kawate, Chiaki Hosoda, Yoichi Kobayashi, Noboru Takayanagi, Tsutomu Yanagisawa

**Affiliations:** Department of Respiratory Medicine, Saitama Cardiovascular and Respiratory Center, Saitama, Japan

## Abstract

**Background:** Coronavirus Disease 2019 (COVID-19) causes severe acute respiratory failure. Antibody-dependent enhancement (ADE) is known as the mechanism for severe forms of other coronavirus diseases. The clinical progression of COVID-19 before and after IgG antibody seroconversion was investigated.

**Methods:** Fifty-three patients with reverse transcriptase PCR (RT-PCT)-confirmed COVID-19 viral pneumonia with or without respiratory failure were retrospectively investigated. The timing of the first IgG antibody against SARS-CoV-2-positive date, as well as changes of C-reactive protein (CRP) as an inflammatory marker and blood lymphocyte numbers, was assessed using serial preserved blood samples.

**Findings:** Ten patients recovered without oxygen therapy (mild/moderate group), 32 patients had hypoxemia and recovered with antiviral drugs (severe/non-ICU group), and 11 patients had severe respiratory failure and were treated in the ICU (6 of them died; critical/ICU group). The first IgG-positive date (day 0) was observed from 5 to 18 days from the onset of disease. At day 0, a CRP peak was observed in the severe and critical groups, whereas there was no synchronized CRP peak on day 0 in the mild/moderate group. In the severe/non-ICU group, the blood lymphocyte number increased (P=0.0007) and CRP decreased (P=0.0007) after day 0, whereas CRP did not decrease and the blood lymphocyte number further decreased (P=0.0370) in the critical/ICU group.

**Interpretation:** The respiratory failure due to COVID-19 viral pneumonia observed in week 2 may be related to an antibody-related mechanism rather than uncontrolled viral replication. In the critical form of COVID-19, inflammation was sustained after IgG seroconversion.

**Funding:** none

## Introduction

Coronavirus Disease 2019 (COVID-19) is a new emerging disease that has affected many countries. Approximately 80% of laboratory confirmed patients have mild to moderate disease, 13.8% have severe disease, and 6.1% have critical disease (respiratory failure, septic shock, and/or multiple organ dysfunction/failure).^1-3^ The pathophysiology of COVID-19 is not well known, but it is currently believed to be primary inflammation initiated by viral replication followed by immune-mediated mechanisms.^4-6^ Similar to SARS-CoV infection, another mechanism facilitating viral cell entry and inflammation, called antibody-dependent enhancement (ADE), has been proposed to account for the severe outcomes in COVID-19.^7-9^ ADE is observed in Dengue fever, Ebola virus, and severe acute respiratory syndrome (SARS) in animal models of the diseases and in clinical practice.^10-13^ The complex of virus and antibody binds to the Fc receptor on macrophages, leading to enhanced viral cell entry/replication and a cytokine response. To date, there has been no clinical evidence about whether IgG seroconversion is related to the rapid worsening of COVID-19 pneumonia.

From February 10, 2020, more than 80 patients with COVID-19 were admitted to our hospital, and acute exacerbations occurred in part of the patients with COVID-19 in week 2 from onset. Therefore, the timing of anti-SARS-CoV-2 antibody seroconversion and the clinical outcomes, especially the changes of C-reactive protein (CRP) and blood lymphocyte numbers, were investigated.

## Patients and Methods

### Patients

Between February 10 and April 30, 2020, 82 patients with COVID-19 admitted to the Saitama Cardiovascular and Respiratory Center were diagnosed by positive polymerase chain reaction with reverse transcription (RT-PCR). All patients underwent high-resolution computed tomography (HRCT), and 74 patients showed viral pneumonia. Viral pneumonia was diagnosed by the consensus of radiologists and respirologists based on the findings of ground-glass appearance and the absence of other findings supporting a different diagnosis. Of the 74 patients, serial preserved blood-test samples could be examined for anti-SARS-CoV-2 IgG antibody in 53 patients. IgG seroconversion was observed during the course in 27 of the patients, and IgG antibody was positive on admission in 26 patients. These 53 patients with COVID-19 pneumonia, composed of a mild/moderate group (no hypoxemia, recovered spontaneously), a severe/non-ICU group (presence of hypoxemia and recovered with nasal/facial oxygen therapy and anti-viral drug therapy), and a critical/ICU group, were studied.

### Methods

The study was approved by the ethics committee of Saitama Cardiovascular and Respiratory Center. The records of daily temperature, oximetry, laboratory findings, radiological changes, and treatments undergone were collected and analysed. Blood tests were done two or three times a week. The first day when the IgG-antibody Kit for SARS-CoV-2 (Kurabo, Ltd., Osaka, Japan) was positive was designated day 0, and the days were then designated in relation to this day as day -6 (±1), day -3 (±1), day 3 (±1), and day 6 (±1).

### Statistical analysis

Data are expressed as means (standard deviation). The values of different groups were compared by the Wilcoxon test or the Kruskal-Wallis test followed by Dunn’s test. Values of day 0 were set as the control value. P<0·05 was considered significant. JMP Ver.13, (SAS Institute Japan Ltd, Tokyo, Japan) was used for statistical analyses.

## Results

Of the 53 patients, 10 had mild/moderate disease (Table 1). All 32 severe/non-ICU patients recovered with anti-viral drugs (favipiravir in 21 patients, lopinavir-ritonavir in 5 patients, hydroxychloroquine in 6 patients) and oxygen therapy. In 15 patients of the severe/non-ICU group, IgG seroconversion was observed during the course in our hospital. In the other 17 patients, SARS-CoV-2 IgG antibody was positive at the time of admission; all of them had been transferred to our hospital because of rapid worsening within 3 days before admission. All 11 critical/ICU patients had severe respiratory failure and were treated with anti-viral drugs (favipiravir in 6 patients, lopinavir-ritonavir in 2 patients, hydroxychloroquine in 2 patients). Of them, 6 died, and 3 were treated by extracorporeal membrane oxygenation (ECMO). In 7 patients in the critical/ICU group, IgG seroconversion was observed during the course in our hospital, and the other 4 patients had been transferred to our hospital because of rapid worsening of the disease, and they were positive for anti-SARS-CoV-2 IgG antibody at the time of admission.

**Table 1.**
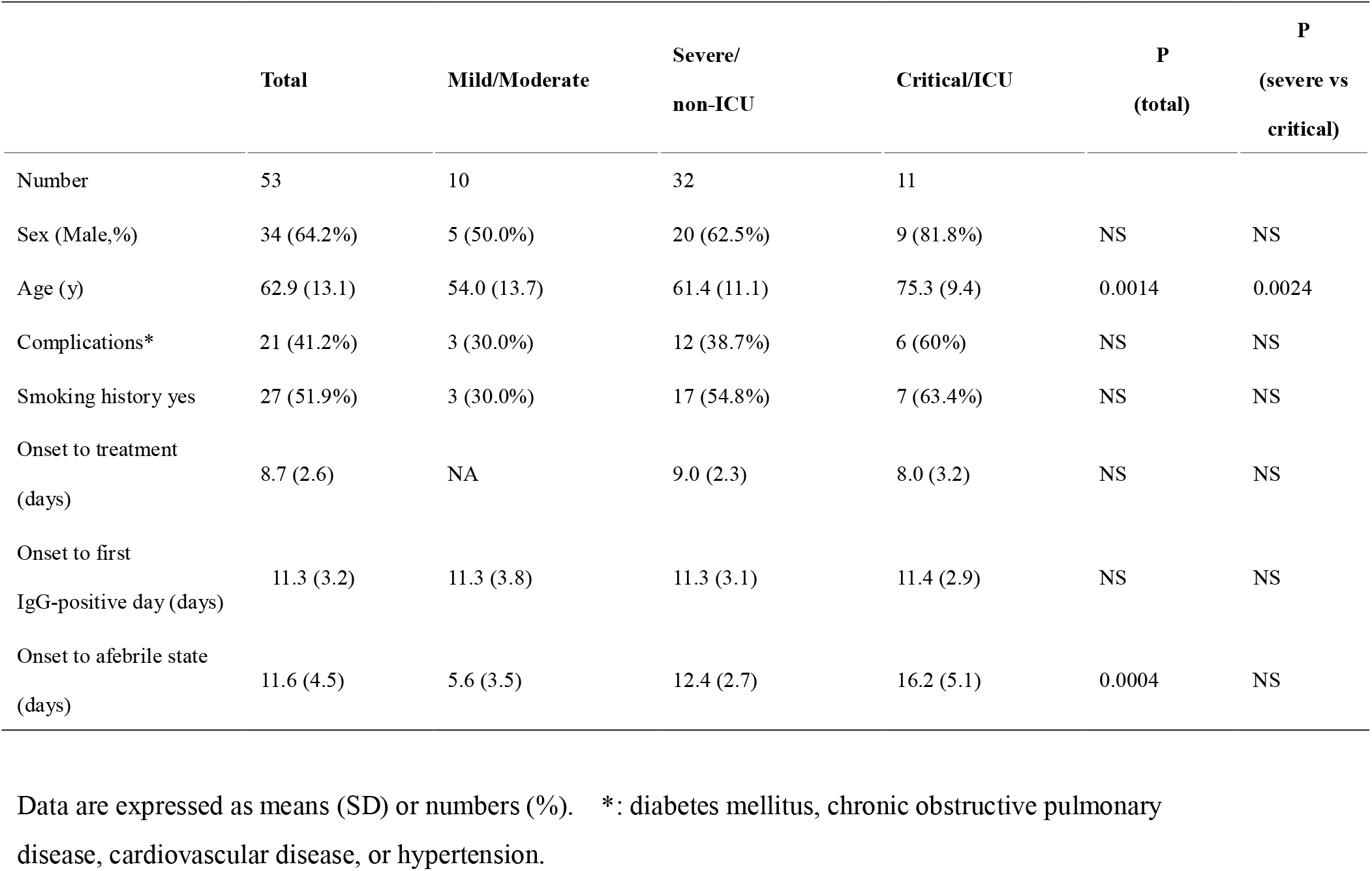
Characteristics of patients with COVID-19 pneumonia

Disease severity increased significantly with older age (P=0.0014). The timing of the start of anti-viral treatment in severe or critical patients was 9 or 8 days after onset, respectively. Days from onset required for IgG seroconversion were not significantly different among the groups (mean 11.3 days).

The blood lymphocyte number on admission was significantly lower in severe or critical patients than in mild/moderate patients (P<0.0001, Table 2). There were no significant differences between severe/non-ICU and critical/ICU patients in D-dimer, LDH, CRP, or ferritin levels on admission.

**Table 2.** Laboratory data on admission

|  | **Total** | **Mild/Moderate** | **Severe/Non-ICU** | **Critical/ICU** | **P**<br>**(total)** | **P**<br>**(severe vs**<br>**critical)** |
| --- | --- | --- | --- | --- | --- | --- |
| PaCO<sub>2</sub> (torr) | 33.5 (4.7) | 38.4 (3.2) | 32.7 (4.2) | 31.3 (4.3) | 0.0011 | NS |
| PaO<sub>2</sub> (torr) | 71.7 (12.3) | 85.1 (11.3) | 69.7 (9.5) | 64.5 (11.6) | 0.0007 | NS |
| WBCs (/μL) | 6203 (2443) | 6920 (1825) | 6159 (2748) | 5681 (1951) | NS | NS |
| Lymphocytes (/μL) | 1088 (521) | 1770 (416) | 984 (413) | 772 (343) | <0.0001 | NS |
| D-dimer (μ/ml) | 2.05 (3.71) | 1.78 (1.97) | 2.21 (4.65) | 1.84 (0.81) | NS | NS |
| AST (IU/L) | 45.5 (31.4) | 30.8 (14.1) | 42.3 (20.1) | 68.0 (53.9) | 0.0337 | NS |
| ALT (IU/L) | 33.5 (23.7) | 35.1 (19.5) | 30.6 (21.5) | 40.4 (32.4) | NS | NS |
| CK (IU/L) | 197.5 (375.4) | 82.5 (60.2) | 154.6 (114.9) | 422.6 (728.1) | 0.0276 | NS |
| LDH (IU/L) | 331.8 (130.9) | 241.8 (94.2) | 328.1 (103.0) | 424.1 (175.3) | 0.0098 | NS |
| CRP (mg/dL) | 6.85 (6.36) | 1.84 (2.61) | 8.21 (7.03) | 7.48 (4.42) | 0.0021 | NS |
| PCT (ng/mL) | 0.10 (0.09) | 0.05 (0.05) | 0.10 (0.10) | 0.12 (0.06) | 0.0161 | NS |
| Ferritin (ng/mL) | 829.4 (675.0) | 435.1 (645.0) | 861.6 (649.2) | 1100.1 (665.3) | 0.0113 | NS |
| KL-6 (U/mL) | 369.2 (343.6) | 298.2 (117.0) | 354.9 (320.9) | 483.0 (525.7) | NS | NS |

Data are expressed as means (SD).
WBCs: white blood cells, AST: aspartate aminotransferase, ALT: alanine aminotransferase, CK: creatine kinase, LDH: lactate dehydrogenase, CRP: C-reactive protein, PCT: procalcitonin.

Figures 1 and 2 show the changes in CRP and blood lymphocyte numbers before and after IgG antibody seroconversion. In patients with mild/moderate viral pneumonia, patients became afebrile 5·6 days from onset (Table 1), and there were no synchronized changes in CRP and blood lymphocyte numbers with IgG seroconversion. In severe/non-ICU patients, CRP increased rapidly from day-3 to day 0 and decreased within 3 days from day 0 (P=0.0001). Blood lymphocyte numbers increased from day 0 in all cases (P=0.0007). In critical/ICU patients, CRP increased rapidly from day -3 to day 0, and it did not decrease significantly to day 3 or day 6. However, blood lymphocyte numbers decreased after day 0 in all cases. Overall, the times of peak CRP values were very close to the times of the first IgG antibody-positive day, rather than the times of the start of treatment (Table 1 and Figure 3).

**Figure 1.**
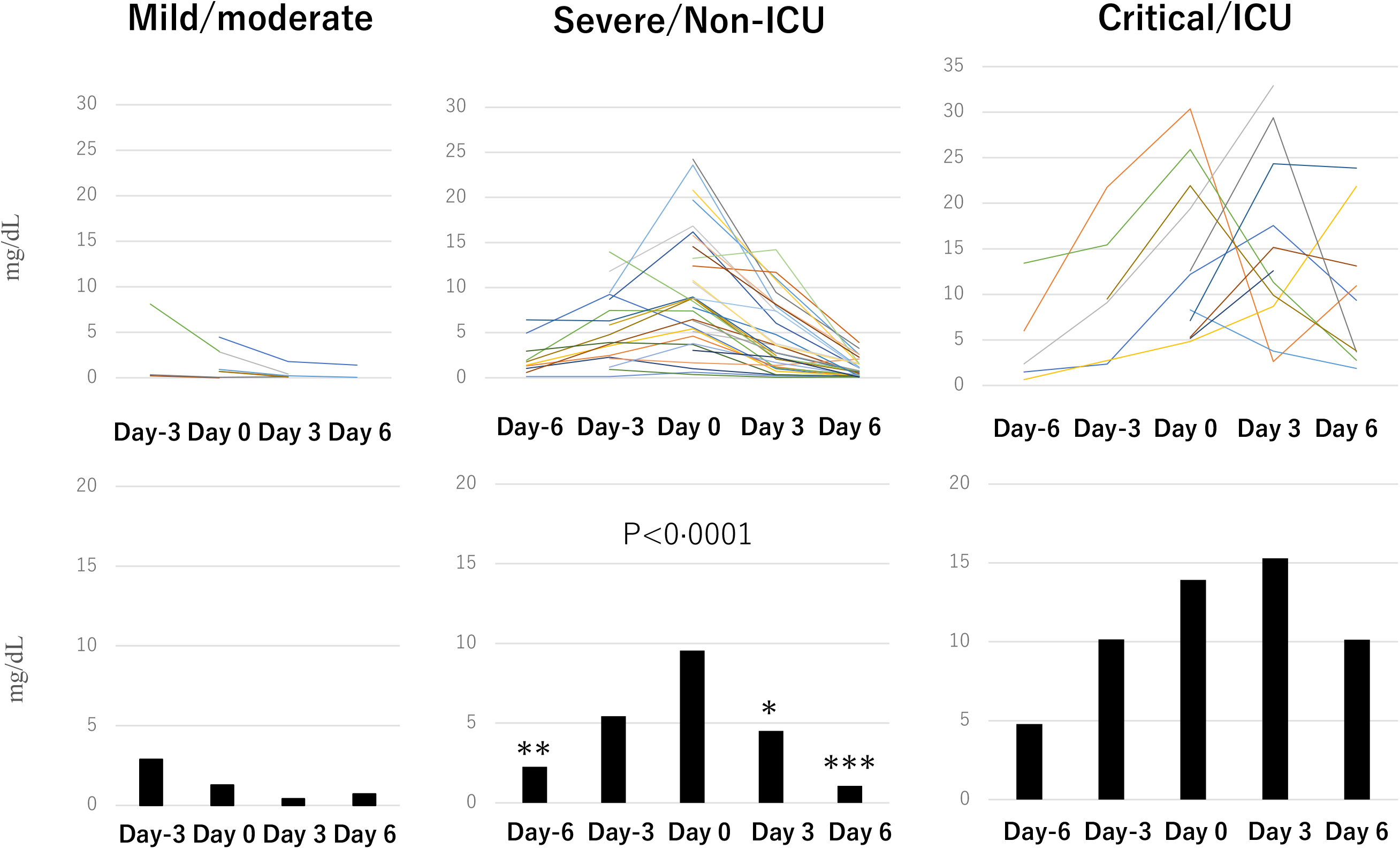
Changes of C-reactive protein before and after IgG antibody seroconversion. The first positive day for IgG-antibody against SARS-CoV-2 is designated day 0, and day -6 (±1), day -3 (±1), day 3 (±1), and day 6 (±1) are designated with respect to day 0. Values of day 0 were set as the control value. * denotes P<0.05, ** denotes P< P<0.01, and *** denotes P<0.001.

**Figure 2.**
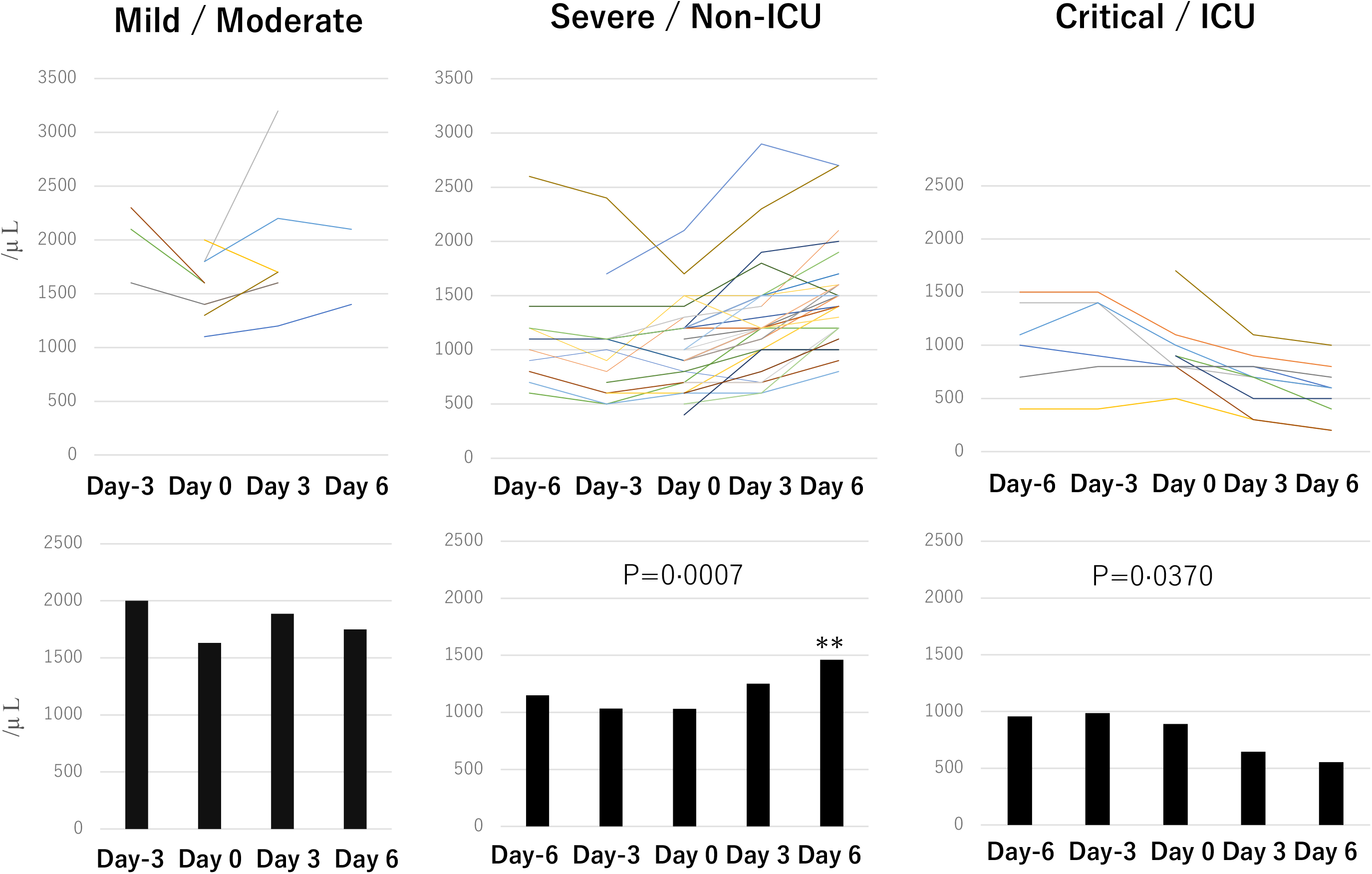
Changes of blood lymphocyte numbers before and after IgG antibody seroconversion. The first positive day for IgG-antibody against SARS-CoV-2 is designated day 0, and day -6 (±1), day -3 (±1), day 3 (±1), and day 6 (±1) are designated with respect to day 0. Values of day 0 were set as the control value. ** denotes P<0.01.

**Figure 3:**
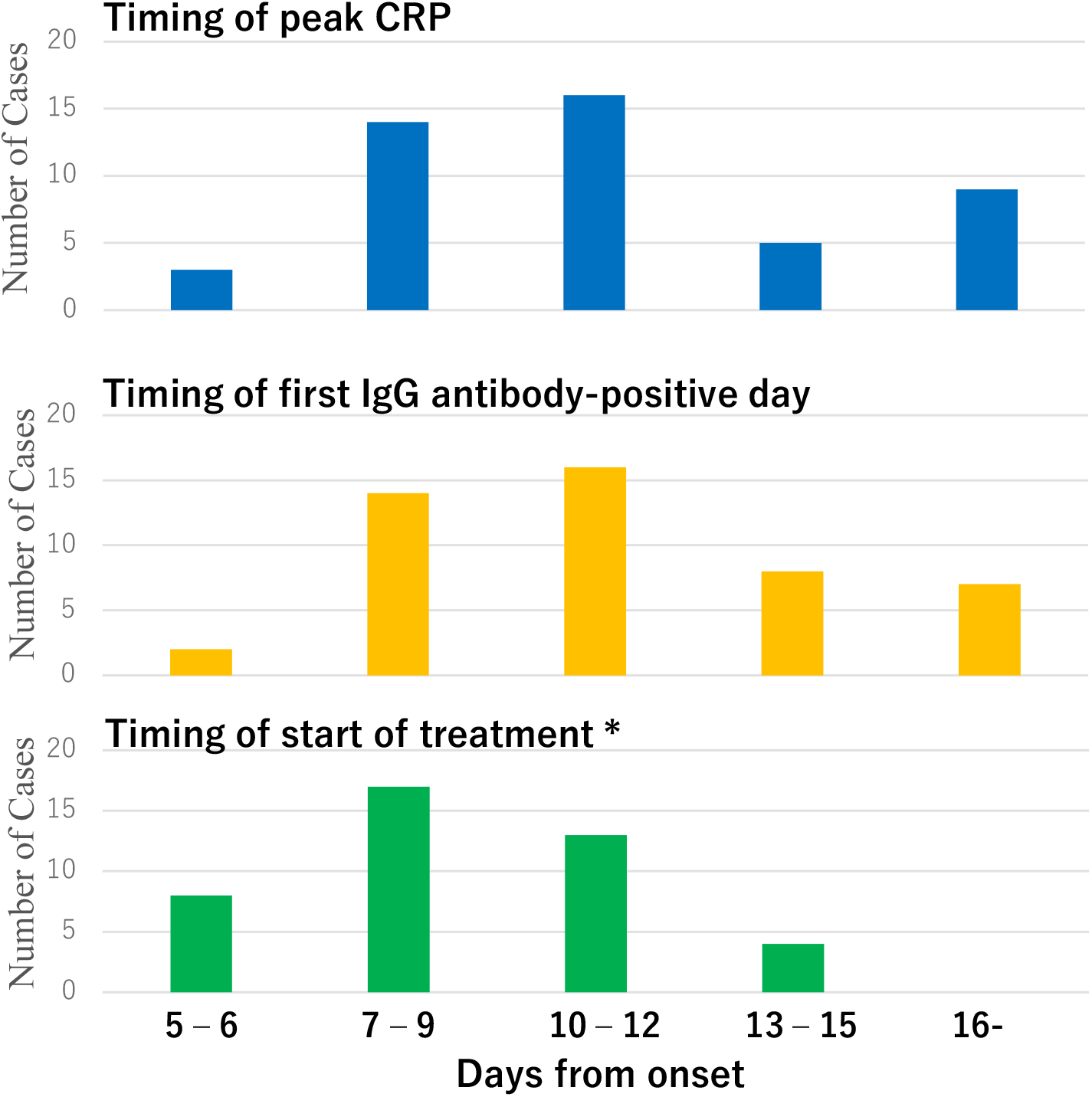
Timings of peak CRP, first IgG antibody-positive day, and start of treatment from onset. *: treatment included favipiravir, lopinavir-ritonavir, or hydroxychloroquine.

## Discussion

This was a retrospective, observational study, but consecutive patients admitted to our hospital were entered, and all patients were screened by HRCT. In addition, all preserved serial blood samples for antibody tests and detailed clinical records could be used, so that it was possible to follow cohorts of patients with COVID-19 pneumonia. The first IgG antibody-positive day was 11 days from onset in all three groups. The results were a little earlier compared to the report from China (median 13 days).^14^ According to the report, IgG antibody starts to increase in week 1. The results of the present study suggest that elevations of the IgG antibody titre to the threshold of the IgG-kit of this study were not significantly different among the groups.

All patients in this study with COVID-19 pneumonia had similar symptoms, such as fever, cough, myalgia, or general malaise in week 1 (data not shown). In the mild/moderate group, these symptoms improved without specific treatments before IgG seroconversion. At this stage, the inflammation was considered the result of viral replication.^6^ Viral replication cause virus-pyptosis, a highly inflammatory form of programmed cell death, as seen in patients with SARS-CoV,^15^ and it is a trigger for the subsequent inflammatory immune response.^16^ In most cases, recruited cells clear the infection in the lung, the immune response recedes, and patients recover.^16^ However, in patients with severe or critical disease, rapid exacerbations of fever, general malaise, hypoxemia, and increased radiological shadows were observed in week 2. The development of respiratory failure coincided with antiviral IgG seroconversion. In nearly half of the patients, the precise IgG seroconversion date could not be determined because they were positive for IgG-antibody on admission. However, all of them had rapid respiratory failure one or two days before they had been transferred to our hospital. After the first IgG antibody-positive day, whether it was during the course in our hospital or on admission, the patients showed quite similar courses of CRP and lymphocyte numbers according to group (non-ICU or ICU groups).

This clinical worsening cannot be explained by uncontrolled viral replication because it is reported that viral load decreases significantly after day 8 from onset.^17^ The findings of the present study suggest that the rapid exacerbation in week 2 is related to the antibody response to SARS-CoV-2. Importantly, there were no significant differences in disease severity as expressed by CRP or PaO2 between the non-ICU and ICU groups at the time of admission. The only difference between the non-ICU and ICU patients on admission was age. It is well accepted that lymphocyte number, D-dimer, and ferritin are predictive markers for fatal COVID-19.^18^ These markers were significantly different in all patients of the present study, but they could not predict non-ICU or ICU patients on admission. The simple immunological response after virus-pyptosis cannot explain why the severities of non-ICU and ICU cases were similar on admission, and why some of them improved quickly and others suffered further disease progression after seroconversion.

Although the present finding is not direct evidence for ADE, the results of the current study can be well explained by the mechanism. ADE of viral entry has been observed in Middle East respiratory syndrome coronavirus (MERS-CoV) and SARS coronavirus (SARS-CoV), coronaviruses very close to SARS-CoV-2. In MERS-CoV ADE, virus entry via the Fc receptor occurs in an antibody dosage-dependent manner.^19^ ADE occurs only at intermediate neutralizing antibody dosages. Another research group observed that higher concentrations of anti-sera against SARS-CoV neutralized SARS-CoV infection, while highly diluted anti-sera significantly increased SARS-CoV infection and induced increased levels of TNF-a, IL-4, and IL-6.^20^ On the other hand, Wu et al. reported that, in patients with mild COVID-19, the titres of neutralizing antibodies were positively correlated with CRP levels on admission.^21^ Taken together, virus-antibody mediated inflammation might be involved in mild disease, but it should be more complicated in the mechanism for severe disease.

In the present study, the increased lymphocyte number after seroconversion in the non-ICU group and the decreased lymphocyte number in the ICU group seemed to be very consistent, and this cannot be explained by simple lymphocyte exhaustion.^22^ In the feline coronavirus (FCoV) ADE model, TNF-alpha is a lymphocyte apoptosis-inducing factor, and its involvement in lymphopenia in FIP cats has been suggested.^23^ One explanation of the present results is that an increase or decrease of lymphocyte number after seroconversion may reflect the termination or worsening of ADE due to the balance of virus load and antibody titre. Further studies of virus load, antibody titre, and the nature of the antibody are needed to answer this question.

There are several limitations that should be noted. First, because of the retrospective nature of the study, not all data points were collected. However, the times of seroconversion were consistent among the groups and with the results of previous reports. In addition, the rapid exacerbations that occurred around day 10 from onset could be traced. Second, the present findings provide only indirect evidence for ADE. Necrotic lymph nodes and atrophic spleens reported in COVID-19 patients^1^ also suggest significant apoptosis of lymphocytes, but more clinical and laboratory data are needed to establish the ADE process in COVID-19. Third, the bias of treatment should be assessed. All patients were treated with anti-viral drugs soon after admission in the severe and critical groups. The drugs were not different between the groups. In addition, the timing of the start of treatment and the timing of peak CRP were not consistent in the two groups.

In conclusion, the results of the present study suggest that severe COVID-19 pneumonia is related to antibody-dependent inflammation. Multiple factors determine whether an antibody neutralizes a virus or causes ADE.^24^ These include the specificity, concentration, affinity, and isotype of the antibody. To elucidate a mechanism for the different clinical courses after seroconversion may help to develop drugs or vaccines against SARS-CoV-2. The safety of and the potentially harmful responses to vaccines to develop ADE antibodies should be carefully assessed in human trials.^25^

## Data Availability

All data are included in the manuscript.

## Authors’ contributions

K Kurashima and K Kagiyama are co-principal investigators, jointly wrote the report, and supervised the clinical component of the study. T Ishiguro, H Nakajima, S Shibata, Y Matsui, K Takano, Y Isono, T Nishida, E Kawate, C Hosoda, Y Kobayashi, T Yotaro, N Takayanagi, and T Yanagisawa were involved in collection and analysis of the clinical data.

## Acknowledgments

We thank all medical and paramedical staffs in Saitama Cardiovascular Center who cared and worked for the patients with COVID-19.

